# NESTApp: A GUI-Based MATLAB Application for Auditable TMS-EEG Preprocessing with EEGLAB

**DOI:** 10.64898/2026.10.11.26365251

**Authors:** Aref Pariz, Wesley Dunne, Jeremie Lefebvre, Sara Tremblay

## Abstract

Electroencephalography (EEG) preprocessing determines signal quality and interpretation, particularly for transcranial magnetic stimulation combined with EEG (TMS-EEG), where recordings are affected by large and complex stimulation-related artifacts. Several preprocessing approaches have been developed, but substantial variability remains in how pipelines are implemented, documented, and evaluated across recordings, limiting reproducibility and comparability across studies. We developed NESTApp, a MATLAB-based graphical interface built on EEGLAB, to provide an accessible and reproducible framework for implementing and evaluating TMS-EEG preprocessing pipelines. Rather than prescribing a specific pipeline, NESTApp allows users to construct and consistently apply customisable workflows incorporating established EEGLAB and TMS-EEG processing tools, while automatically documenting processing decisions and parameters. Integrated quality-control procedures track data removal and flag recordings based on user-defined criteria, facilitating systematic evaluation across participants. The application also supports group-level visualisation and export of processed TMS-EEG measures for subsequent analyses. Applied to 29 recordings from 16 participants, NESTApp processed 27 and identified those requiring further inspection. By integrating pipeline implementation, documentation, quality control, and group-level evaluation within a single interface, NESTApp provides a transparent framework for TMS-EEG preprocessing that may improve reproducibility, facilitate consistent processing across cohorts, and lower technical barriers to the implementation of established preprocessing approaches.

## Introduction

Combining transcranial magnetic stimulation (TMS) (Barker et al., 1985) with electroencephalography (EEG) measures cortical responses to stimulation across a wide range of brain regions (Casarotto et al., 2010; Lioumis et al., 2009; Rogasch and Fitzgerald, 2013). Over nearly three decades, TMS-EEG has become an established approach for probing cortical excitability, oscillatory dynamics, and effective connectivity, deepening understanding of brain network organisation and supporting individualised therapeutic protocols in psychiatry and neurology (Esposito et al., 2020; Hui et al., 2019; Saari et al., 2018; Tremblay et al., 2019). TMS-EEG recordings are also among the most heavily contaminated in electrophysiology: on top of the ocular, muscular, environmental, and cardiac artifacts any EEG carries (Sazgar and Young, 2019), the pulse adds coil discharge and magnetic field interactions (Hernandez-Pavon et al., 2023; Ilmoniemi et al., 2015), cranial muscle activity (Mutanen et al., 2013), evoked blinks (Corthout et al., 2011; Sohn et al., 2004), decay and recharge artifacts (Veniero et al., 2009), electrode movement (Gorjan et al., 2022), and ringing (Rogasch et al., 2013).

EEGLAB (Delorme and Makeig, 2004) is a MATLAB toolbox widely used for EEG processing, but applying the same analysis to a set of recordings generally requires scripting, which could be an obstacle for users without a programming background. A second constraint is independent of the first. The methods written for TMS-EEG artifacts are more often published as whole pipelines than as components: the ordering, the parameters and the individual operations arrive together, so adopting one step means adopting the sequence around it, and a method well suited to a dataset cannot easily be lifted out and placed in a different order. A method also stays available only while the pipeline carrying it is maintained. TESA (TMS-EEG Signal Analyzer) (Mutanen et al., 2020; Rogasch et al., 2017) is a notable exception. It integrates with EEGLAB and provides separate functions for decay removal, TMS-locked epoching, and independent component analysis (ICA), each usable in an order of the user’s choosing. Distributed this way, a method can be combined with others and outlives the pipeline it came from; distributed whole, it can do neither.

Script-based work motivated graphical programs such as TMSEEG (Atluri et al., 2016), which lower the coding barrier by fixing the sequence instead. But pipelines vary with the data collection protocol and with the artifacts present, and a systematic comparison shows that such methodological choices change the resulting TMS-evoked potentials (Beck et al., 2024).

The main challenge is to apply the same pipeline consistently across all recordings in a cohort and verify that it works as intended for each recording. We built NESTApp, a GUI in which the user chooses tools available as EEGLAB plugins and applies the saved pipeline to a cohort of resting-state or TMS-EEG recordings without scripting. The *pipeline designer*, typically a principal investigator or senior trainee, selects and orders preprocessing steps, configures their parameters, and validates the workflow on representative datasets, which requires familiarity with EEGLAB and the underlying signal-processing methods. The *pipeline operator*, who could be a junior trainee, research assistant, or collaborator, loads a saved pipeline, selects data files, and presses a button; the role needs no scripting. The application meanwhile documents each step, validates software dependencies before the run, and writes a quality report for each file that the designer can check afterwards.

Sharing analysis code is an important step toward reproducible EEG preprocessing and is increasingly encouraged. However, code primarily documents the intended processing workflow; the outcome of that workflow may differ across recordings depending on data quality, artifact removal, excluded channels or trials, and other data-dependent decisions. Maintaining a complete record of these processing outcomes therefore requires systematic documentation at the level of each recording. Although this can be implemented within existing analysis environments, it often requires additional coding and may be inconsistently applied, particularly when preprocessing is conducted across large cohorts or by multiple researchers.

NESTApp was developed to address this gap by integrating pipeline implementation and documentation within the same framework. Preprocessing pipelines are stored as structured, portable configurations rather than embedded within analysis code, allowing the same workflow to be saved, shared, modified, and consistently applied across recordings and users. For each recording, NESTApp automatically documents the processing steps, parameter values, and data-quality decisions that contributed to the final cleaned dataset. At the cohort level, this information is integrated into standardised reports and a methods description reflecting the preprocessing that was performed. This approach provides a transparent and traceable link between the intended preprocessing pipeline, its application to individual recordings, and the methods ultimately reported, facilitating reproducibility across researchers and datasets.

### Related Work

Existing EEG analysis environments differ in how far they support flexible pipeline construction, graphical operation, and batch processing across recordings. FieldTrip (Oostenveld et al., 2011), MNE-Python (Gramfort et al., 2013), and BeMoBIL (Klug et al., 2022) impose no constraints on how a pipeline is constructed, because a pipeline in these environments is written as code. This flexibility also means that both building and running a pipeline require programming experience. EEGLAB (Delorme and Makeig, 2004) offers a GUI for interactive, function-by-function processing and can record the steps performed as a script, but applying the same sequence to new files means editing and re-running that script by hand. Brainstorm (Tadel et al., 2011) executes batches through its Process tab, and its pipeline editor saves a pipeline for reuse or exports it as a MATLAB script. HAPPE (Gabard-Durnam et al., 2018) trades flexibility for automation in developmental and high-artifact EEG, applying a standardised automated pipeline and generating quality metrics without being asked; it is configured by script, covers resting-state and event-related data, and provides no steps for TMS artifacts.

TMS-EEG preprocessing is commonly done by scripting EEGLAB together with TESA (Mutanen et al., 2020; Rogasch et al., 2017), which supplies the TMS-specific operations EEGLAB lacks. That combination places no limit on what a pipeline can do, but the order, the parameters, and the application of the sequence to each recording are the user’s own code, and so is any account of what happened to each file. Among the tools that package a sequence instead, TMSEEG (Atluri et al., 2016) presents a step-by-step GUI over a fixed order, while ARTIST (Wu et al., 2018) and AARATEP (Cline et al., 2021) run automatically from scripts with fixed step orderings.

These tools use a fixed processing sequence, which is a reasonable way to reduce pipeline variability. However, this approach has three limitations. A pipeline validated on one protocol, montage, and population is not thereby correct for another, so the choices are standardised without being made appropriate to the data in hand. Standardisation also holds only among the users of each pipeline, and several exist, which relocates the variability between laboratories rather than removing it. Above all, an advance made in one pipeline cannot be added to another. For example, ARTIST classifies independent components using a Fisher linear discriminant trained on expert-labelled components (Wu et al., 2018), whereas AARATEP uses autoregressive-blended reconstruction of the peri-pulse window (Cline et al., 2021). These methods address different problems and could, in principle, be applied to the same recording. They are distributed as parts of separate pipelines rather than as independent processing steps, so advances that might accumulate remain alternatives.

Table 1 compares NESTApp and the tools above across workflow infrastructure features. Two of those features matter together rather than separately: a pipeline that can be handed to someone else, and batch execution that does not require them to write code. EEGLAB itself provides both, through the command history it records and through its STUDY interface. That history is a log of the calls issued, so it records what was asked for rather than what resulted: it does not report that one channel was interpolated in one recording and thirteen in another, nor summarise such figures across a cohort. Holding the pipeline as data rather than as code is what lets NESTApp compose steps from different sources, hand the result to someone else, and report the outcome for every file.

**Table 1.** Workflow features of the EEG preprocessing tools cited in this work.

| Tool | GUI | Custom step ordering | Shareable pipeline | Batch execution | Dependency check | Auto QC reports | Dedicated TMS-EEG steps | Resting EEG | Runs without a paid software licence |
| --- | --- | --- | --- | --- | --- | --- | --- | --- | --- |
| FieldTrip | – | ✓<sup>a</sup> | ✓<sup>a</sup> | ✓<sup>a</sup> | – | – | ✓ | ✓ | – |
| Brainstorm | ✓ | ✓ | ✓<sup>b</sup> | ✓<sup>c</sup> | – | – | – | ✓ | ✓<sup>d</sup> |
| MNE-Python | – | ✓<sup>a</sup> | ✓<sup>a</sup> | ✓<sup>a</sup> | – | –<sup>e</sup> | ✓ | ✓ | ✓ |
| EEGLAB | ✓ | ✓<sup>f</sup> | ✓<sup>a</sup> | ✓<sup>g</sup> | ✓<sup>h</sup> | – | ✓<sup>i</sup> | ✓ | ✓<sup>d</sup> |
| TESA | ✓<sup>j</sup> | ✓ | ✓<sup>a</sup> | ✓<sup>a</sup> | – | – | ✓ | – | – |
| TMSEEG | ✓ | – | – | – | – | – | ✓ | – | – |
| ARTIST | – | – | ✓<sup>a</sup> | ✓<sup>a</sup> | – | – | ✓<sup>k</sup> | – | – |
| AARATEP | – | – | ✓<sup>a</sup> | ✓<sup>a</sup> | – | – | ✓ | – | – |
| HAPPE | – | – | ✓<sup>l</sup> | ✓<sup>a</sup> | – | ✓ | – | ✓ | – |
| BeMoBIL | – | ✓<sup>a</sup> | ✓<sup>a</sup> | ✓<sup>a</sup> | – | ✓ | – | ✓ | – |
| NESTApp | ✓ | ✓ | ✓ | ✓ | ✓ | ✓ | ✓ | ✓ | –<sup>m</sup> |

✓ = supported; – = not supported or not applicable. Checked against each tool’s current documentation in September 2026. Custom step ordering: the user can choose and order preprocessing steps. Shareable pipeline: the configuration can be saved and given to someone else. Dependency check: a missing plugin or toolbox is reported rather than failing at the point of use. Auto QC reports: structured per-file summaries are produced without being asked for. Dedicated TMS-EEG steps: the tool provides routines written for TMS artifacts.
<sup>a</sup>Expressed as code, so sharing or reordering means editing a script. <sup>b</sup>Through Brainstorm’s pipeline editor, or exported as a MATLAB script. <sup>c</sup>Through Brainstorm’s Process tab, within its database. <sup>d</sup>A compiled distribution running on the free MATLAB Runtime. <sup>e</sup>A reporting facility exists, but the user’s script writes the summary. <sup>f</sup>Calls made in the GUI, recorded as a script to edit and re-run. <sup>g</sup>Multiple datasets, through the STUDY interface. <sup>h</sup>The MATLAB toolbox licences are tested at startup; a plugin’s own dependency is not, and fails when its step runs. <sup>i</sup>Through plugins or external toolboxes. <sup>j</sup>Dialogs from EEGLAB’s menus, one per function.
<sup>k</sup>Single-pulse TMS-EEG. <sup>l</sup>Saved and reloaded as a set of parameters, entered at the command line. <sup>m</sup>Requires MATLAB R2023b or later with the Signal Processing Toolbox and the Statistics and Machine Learning Toolbox; the Parallel Computing Toolbox is needed only for parallel batch execution.

### Application Design and Features

#### Architecture

NESTApp is a thin layer over EEGLAB: most pipeline steps are a single EEGLAB or plugin function call, such as filtering, ICA, or artifact rejection, and the interface is a MATLAB App Designer class with three tabs, *Cleaning, Reports*, and *Explore*, in the order a user meets them. A pipeline is an ordered list of steps, each with a name and a structure of typed parameters (Fig. 1). The interface and the execution engine use the same pipeline representation, and a pipeline built through the interface can also be run by calling the engine directly from a script. The same pipeline can therefore run on another machine, or for another user, without opening the application.

**Figure 1.**
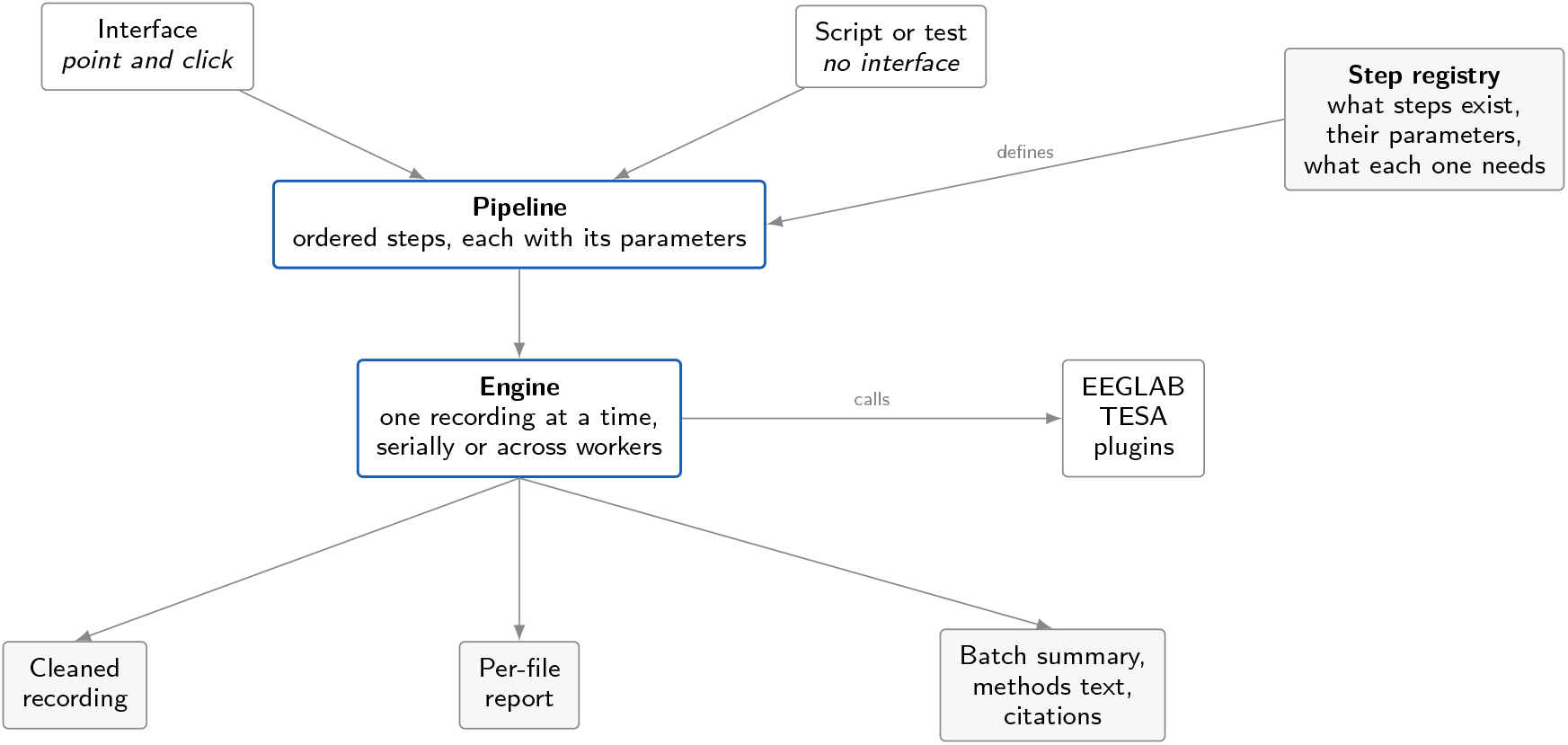
How a run is put together. A pipeline is an ordered list of steps, each carrying its own parameters, and it is the object the user actually builds. The interface is one way to build it and a script is another; both hand the same pipeline to the same engine, which applies it to one recording at a time, serially or distributed across workers, calling EEGLAB, TESA, and the installed plugins to do the work. A single registry declares what steps exist, what parameters each takes, and which plugin each depends on, and every part of the application reads from it, which is what keeps the step list, the dependency check, and the parameter panel in agreement. Each run leaves the cleaned recording, a report for every file, and a batch summary that includes a methods paragraph and the citations for the methods that ran.

A single registry declares what steps exist, what parameters each takes, their defaults, units, valid ranges, and which plugin each depends on. Every part of the application reads from it, which is what keeps the parts consistent: the step list offers exactly the steps whose dependencies resolve on that machine, the pre-flight check tests the same requirements, the installation diagnostic reports from the same declarations, and the parameter panel builds itself from the same entries. Adding a step is therefore one registry entry and one execution case, with no change to the interface code. An automated test dispatches every registered step to confirm that each one offered is implemented. Table 2 counts the steps each processing stage offers and the toolboxes that supply them. Supplementary Table S1 names every step individually.

**Table 2.** Preprocessing steps available in NESTApp, by processing stage.

| Stage | Steps | Provided by |
| --- | --- | --- |
| Import & Session | 4 | EEGLAB |
| Channels | 7 | EEGLAB, clean_rawdata, TESA |
| TMS Artifact | 11 | TESA, AARATEP |
| Reference & Resample | 2 | EEGLAB |
| Detrend | 4 | EEGLAB, TESA |
| Filter | 5 | firfilt, TESA, CleanLine |
| Automatic Cleaning | 3 | clean_rawdata, EEGLAB |
| Epochs & Trials | 4 | EEGLAB |
| ICA | 8 | FastICA, EEGLAB, PICARD, TESA, ICLabel |
| TEP Analysis | 3 | TESA |
| Utilities & Inspection | 3 | nestapp, EEGLAB |

The registry also settles what a given installation can offer, and the dependencies behind that fall into three tiers. NESTApp will not start without MATLAB R2023b or later, the Signal Processing Toolbox, the Statistics and Machine Learning Toolbox, and EEGLAB; the version described here was developed against MATLAB R2026a and EEGLAB 2026.0.0, and runs with EEGLAB 2025 releases. Seven EEGLAB plugins, all installable through its extension manager, supply the steps the built-in templates use: TESA (Rogasch et al., 2017), FastICA (Hyvarinen, 1999), PICARD (Ablin et al., 2018), ICLabel (Pion-Tonachini et al., 2019), CleanLine (Mullen, 2012), `clean_rawdata` (Kothe and Makeig, 2013), and `firfilt` (Widmann et al., 2015). The remaining dependencies are optional, and each enables a single capability: the Parallel Computing Toolbox enables parallel batch execution, the Curve Fitting Toolbox supports a decay-removal step required by AARATEP, and three reader plugins add support for the BrainVision, Neuroscan, and Curry file formats.

Since a step whose dependency is absent is not offered, what a user has to install follows from the pipeline they intend to build rather than being settled in advance. A *Preferences* dialog holds settings that persist between sessions, among them the EEGLAB installation path and a cap on parallel workers. The path setting puts EEGLAB and its plugins on the path at each launch, so the user maintains no startup file; the worker cap is disabled, with the reason given, when the Parallel Computing Toolbox is not licensed.

Before any file is opened, a pre-flight check resolves every step against the installed plugins. A run in progress can be cancelled. *Parallel processing* distributes files across MATLAB workers. Each worker is limited to a share of the machine’s computational threads, because MATLAB otherwise grants every worker as many threads as there are cores, and they oversubscribe the processor. EEGLAB is initialised once per worker rather than once per file, and `ALLEEG` holds only the dataset of the file in hand, so no run can pick up another’s data. Parallelism is requested rather than guaranteed: the run falls back to serial, and states the reason, when one file is selected, when the Parallel Computing Toolbox is not licensed, or when a step in the pipeline opens a dialog and cannot run on a worker.

#### Cleaning

A pipeline is assembled in the *Cleaning* tab (Fig. 2) and saved in MAT format for later batch processing or for sharing. Each catalogue entry names the underlying EEGLAB function and gives practical guidance, for example “Recommended: run after epoching so a pre-stimulus period exists”. For advanced users, *Manual Command* takes MATLAB code of the form `EEG = someFunction(EEG`, arguments), so any function on the path can run as a pipeline step, whether from a toolbox NESTApp does not catalogue or from a file the user wrote. A method too new or too specific to have been packaged can therefore run in a batch, in a chosen position. The application cannot check what such a step does, so it is exempt from the ordering rules. The methods text records that a custom step ran without quoting it, and the command itself is kept in the saved pipeline. A *Quality Gate* is a step of a third kind. Placed at a chosen position, it compares the metrics reached at that point with thresholds set on the step and records the recording as passing, marginal, or failing there, without altering the data. A pipeline that does not end with a *Save New Set* step discards its own work; that step either appends a suffix to the original filename or replaces it with a new name.

**Figure 2.**
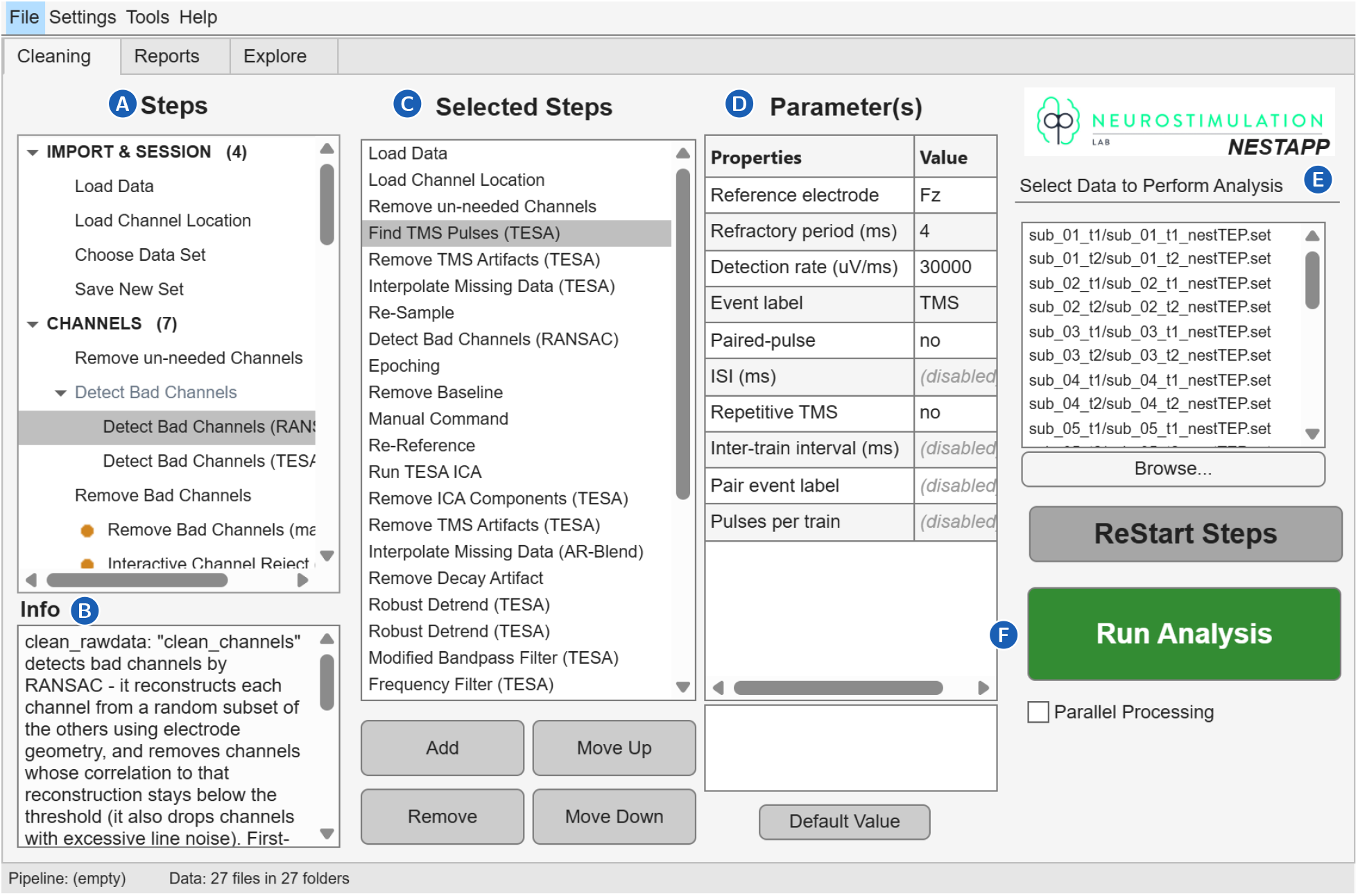
Building and running a pipeline in the *Cleaning* tab. **A** The step catalogue, grouped by processing stage with the number of steps in each. A step whose plugin is absent is not listed, so a pipeline cannot be built around one that will not run. **B** The description of the step selected in the catalogue. For the RANSAC detector shown, it gives the method and the correlation below which a channel is dropped. **C** The pipeline being assembled, here 32 steps. With the parameter values at D, this is what a saved pipeline file contains. **D** The parameters of the step selected in C, here TMS pulse detection: Fz as the reference electrode, a 4 ms refractory period, a 30,000 *μ*V/ms rate of change. **E** The 29 recordings chosen for the run, across 16 participant folders. **F** *Run Analysis* and the parallel-processing checkbox. B and D follow different selections: the description tracks the catalogue; the parameters track the pipeline.

A user can also load one of four pipeline templates that ship with NESTApp. Three populate the pipeline with individual steps to inspect, reorder, or modify: a two-round ICA TMS-EEG pipeline following Rogasch et al. (2017), a resting-state EEG pipeline based on the PREP pipeline (Bigdely-Shamlo et al., 2015), and a minimal event-related potential pipeline following Delorme (Delorme, 2023). The fourth, the AARATEP pipeline (Cline et al., 2021), is instead a single orchestrating step that runs the published pipeline whole, because reproducing the decomposition step by step would make NESTApp responsible for the fidelity of a pipeline it does not own, and any drift would be silent: a mis-ordered pipeline still produces plausible output. Its helper functions are installed on request at a pinned release rather than copied into NESTApp’s source, which keeps the two projects separate and leaves the pipeline its authors’ code at a version the user can name. Three of its steps are nonetheless offered individually, so its pulse detection, its autoregressive-blended interpolation, and its decay fitting can each be placed in a pipeline of the user’s own ordering. Further published pipelines can be added the same way.

Once a pipeline has been selected or built, its steps need to be configured. The parameter panel uses the registry to display human-readable parameter names and units: it shows “High-pass cutoff (Hz)” instead of the raw EEGLAB key `high`. A parameter is disabled if it conflicts with a selected alternative, or if another parameter has not been set to the value required to enable it. Missing values for required parameters without suitable defaults, such as the epoch window, are flagged when a template is loaded, before batch processing begins.

#### Reports

A pipeline report is generated for each dataset processed through the *Cleaning* tab. The report documents how the recording was processed, so a run can be audited without reopening the data. Pipelines that run more than one round of ICA (Rogasch et al., 2017) are reported round by round. When two or more files are processed in a single batch, a cross-file summary is prepended to the report list, giving mean ± SD and range for each metric across the batch. A quality dashboard summarises those metrics against the limits set for them (Fig. 3). Reports from previous sessions load from disk and appear in the same list as the current session. The *Export Metrics CSV* button writes a spreadsheet of key metrics across all visible reports.

**Figure 3.**
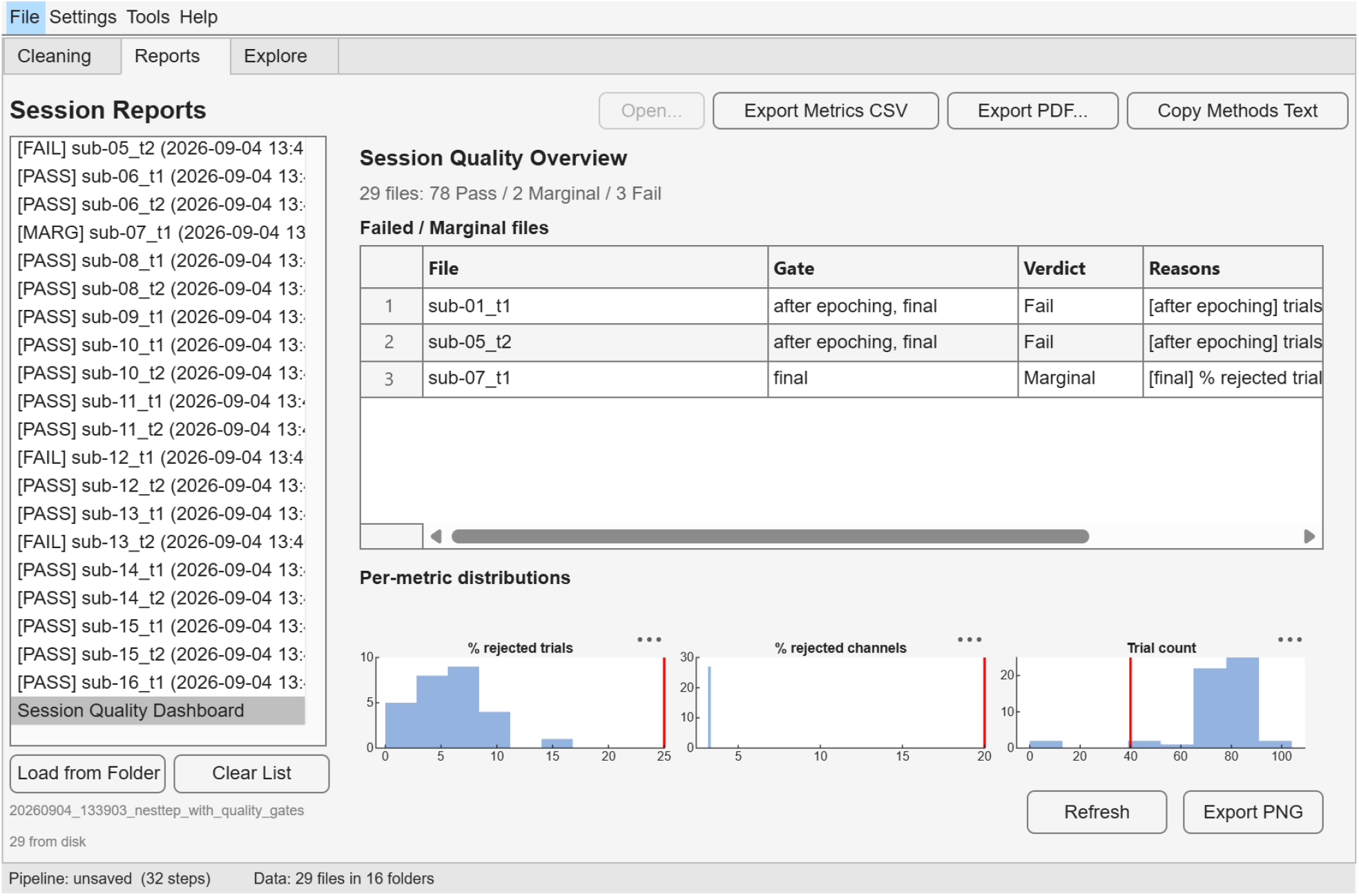
The session quality dashboard. Verdicts for a 29-recording batch. Counts are gate verdicts: three from each of the 27 completed recordings, and one each from the two that stopped at TMS pulse detection, which had cleared the gate at load before stopping. The table names every recording that failed or was marginal, with the gate and the reason it gave, and the histograms below place each metric against the threshold set for it, drawn in red. The list at the left carries a verdict beside every recording, so the files needing attention are apparent without opening any of them. That verdict is the run’s outcome rather than a gate’s: the two recordings that stopped at TMS pulse detection are marked failed there although the one gate they reached had passed, which is why the table above lists neither.

Table 3 states what a run leaves behind, and where. The saved pipeline file holds the step list, the parameters, and a schema version, narrow by design so that it stays portable and can be migrated forward when a step is renamed. The report holds what that pipeline did to one recording. For an ICA step, the pipeline file records that components classified as ocular by ICLabel were to be removed at an ICLabel probability of 0.9 or above; the report records that one such component was identified in one recording, accounting for 1.2% of the variance there, and a different proportion in every other file.

**Table 3.** What a batch run records, and where.

| Output | Contents |
| --- | --- |
| `<name>.set` | The cleaned dataset. Its `EEG.history` holds the full step list with every parameter and a timestamp |
| `<name>_report.mat` | The input filename and the time it was processed; channels original, rejected, interpolated, and final, by name; trials original, rejected, and final; independent components per round with assigned category and variance removed; elapsed time and channel or trial change for every step; *Quality Gate* verdicts where thresholds were set |
| `<name>_report.pdf` | The same report as a document, when enabled |
| `_batch/spec.mat` | The executed pipeline and its name, reloadable into the application |
| `_batch/session_summary.csv` | One row per file: status, step count, duration, quality verdict, failing step and reason, and the channel, trial, and component counts |
| `_batch/session_summary.txt` | Cross-file mean, standard deviation, and range for each metric; the step list; a generated methods paragraph; and citations with DOIs for the methods that ran |
| `_batch/failed_files.txt` | Inputs that did not complete, as a re-run list |
| `_batch/dashboard.png` | Batch quality overview |
| `qc/<name>/` | Checkpoint quality images, when enabled |
| *Partly recorded* | The methods text names the application version; MATLAB, EEGLAB, and the plugins are named but not versioned |

The application also writes the methods prose for the batch (Fig. 4). Of everything a run records, that text is the part that reaches a reader of a published result rather than only whoever holds the dataset. It says what the pipeline did, not whether the pipeline was the right one.

**Figure 4.**
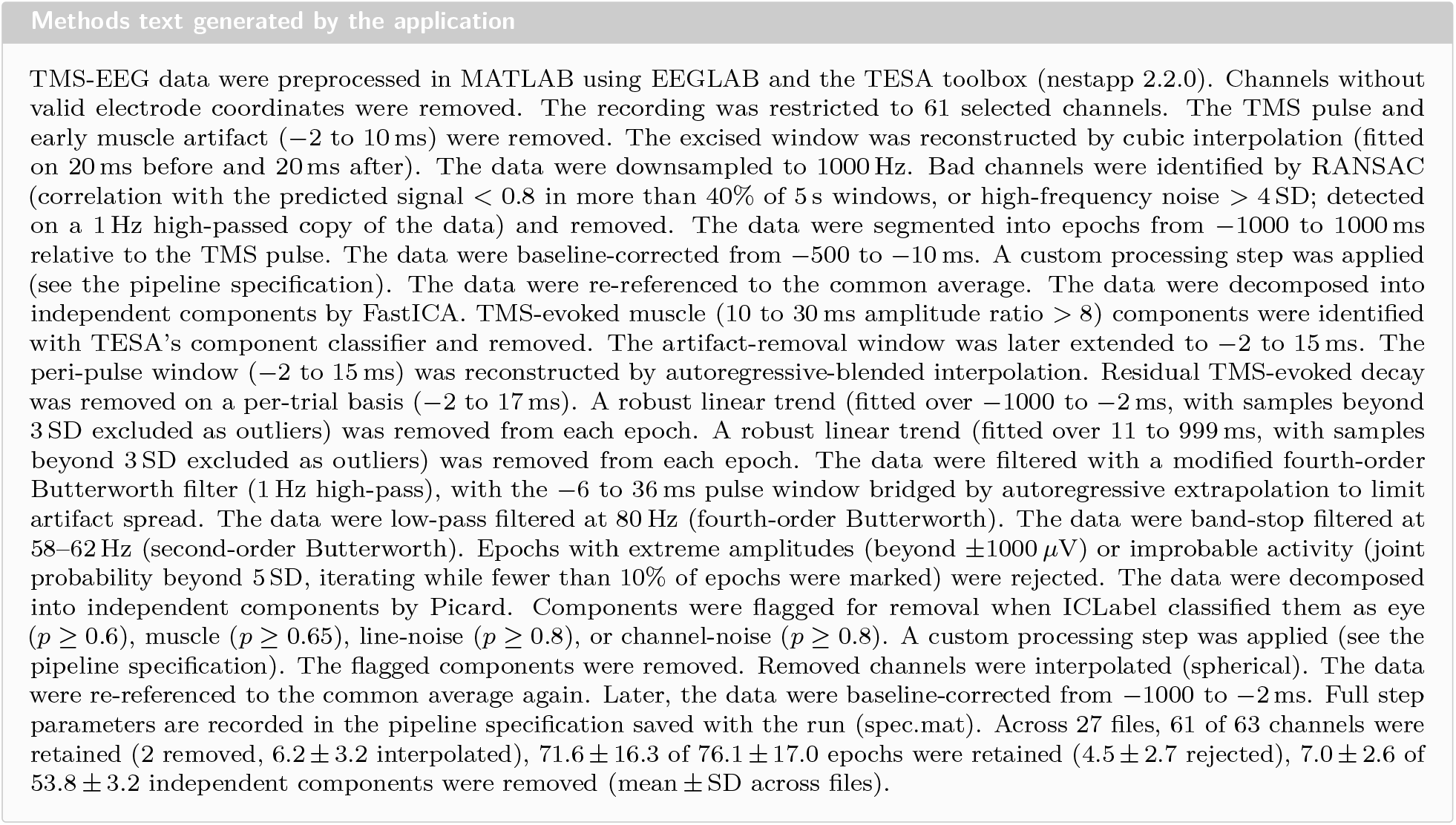
Methods prose written by the application, for the batch reported here. Reproduced without editing. Every clause is emitted by a step that actually executed and carries the values that step ran with, so the paragraph is complete, in that no step that touched the data is silently absent, and specific, in that the windows, cutoffs, and filter orders it quotes are the values used rather than the defaults they were chosen from. A step reordered, reconfigured, or removed changes the sentence that reports it. The counts closing the paragraph are taken from the reports of the files described.

#### Explore

The *Explore* tab supports the comparison most TMS-EEG studies are designed around: pre- versus post-stimulation sessions, or one cohort against another. Cleaned recordings are assigned to any number of named groups (Fig. 5), either by filtering on filename patterns or by editing a table of files, subjects, and group membership. A group is defined over subjects rather than files, so where one participant contributed more than one recording to the same group, those recordings are averaged before the group estimate is formed, and the reported sample size counts participants. Recordings assigned to different groups are never averaged with each other. Averaging files instead would treat four sessions of one participant as four independent observations, inflating the sample size and narrowing the interval. A subject’s mean weights each session by its trial count, because two sessions of one person are not equal evidence when one held 10 trials and the other 190. Unpaired comparisons take the ordinary confidence interval about each group mean, formed across subjects. Paired designs use complete cases, with Cousineau–Morey intervals for within-subject data (Cousineau, 2005; Morey, 2008).

**Figure 5.**
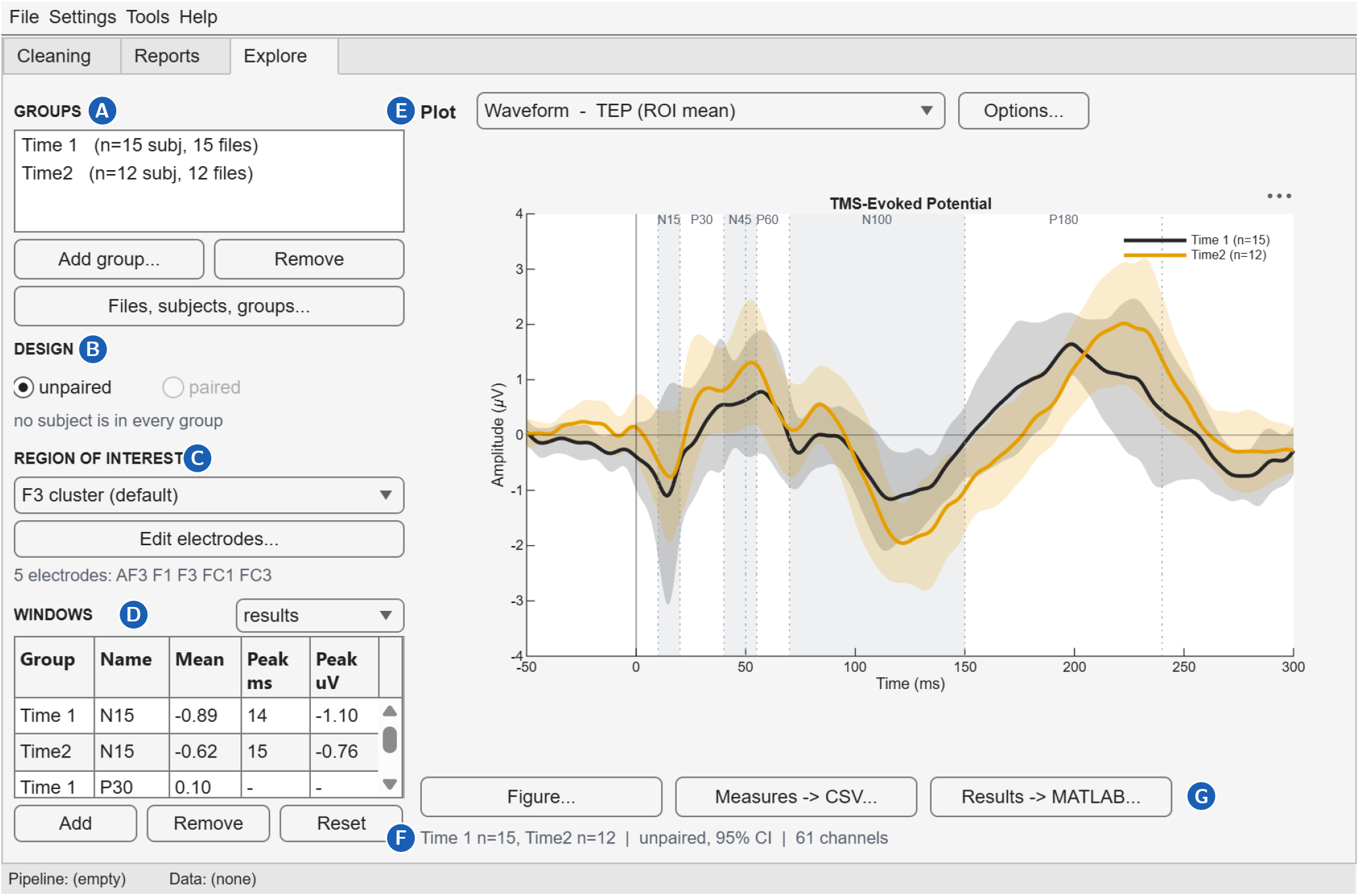
Defining a comparison in the *Explore* tab. **A** The groups, here the two recording sessions, with 15 recordings assigned to Time 1 and 12 to Time 2. **B** The design, here an unpaired comparison. **C** The region of interest, here the default F3 cluster, resolving to the five electrodes named below the selector. **D** The windows, in results mode. N15 gives a mean of −0.89 *μ*V and a peak of − 1.10 *μ*V at 14 ms; P30 and N45 give a mean but no peak. **E** The view drawn, one of the six in Fig. 6. **F** The status line: sample sizes, design, confidence level, and channel count. **G** The three exits, which differ in what they preserve. *Figure* composes an image at a chosen column width and resolution, with the groups, sample sizes, design, and region of interest stamped into it. *Measures* writes one row per participant, group, and window, for statistics done elsewhere. *Results* writes the whole analysis, curves included, and reopens through *File → Load Analysis* so it can be resumed or handed to someone else.

The main area draws a catalogue of plots from that one set of settings (Fig. 6). A plot that cannot be drawn with the current settings, such as a difference wave when three groups are defined, is listed together with the reason. Plot-specific settings are reached through an options dialog generated from the same registry that defines the plots.

**Figure 6.**
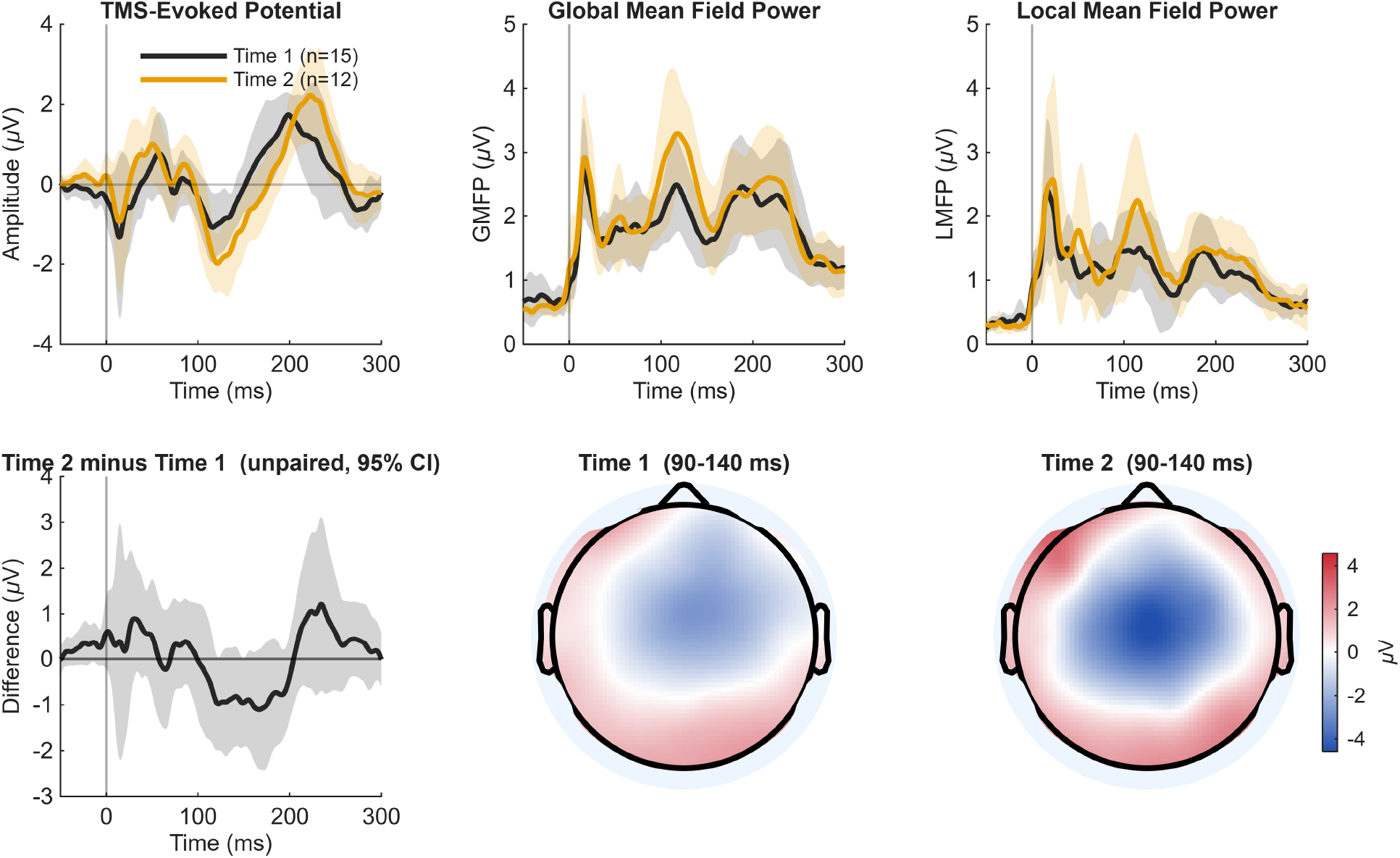
The views the *Explore* tab offers, over one cohort. Twenty-seven cleaned recordings from 16 participants, assigned to two groups by the session they came from. **Top row:** the region-of-interest waveform, global mean field power, and local mean field power over the region of interest. Shaded bands are 95% confidence intervals formed across participants. **Bottom row:** the difference between the groups, and the scalp distribution of each group averaged over 90 to 140 ms on a shared colour scale. A group is defined over subjects rather than files, so repeated recordings of one participant are averaged before the group estimate is formed and the reported *n* counts participants; sessions are weighted by their trial counts. All six panels are drawn from one set of settings, so changing the region of interest or the window redraws every view that depends on it. The figure was composed by the application’s own publication export at the width it is printed here, rather than captured from the screen.

The estimates are descriptive: group means with confidence intervals and the corresponding per-window measures. They let an investigator see the size and direction of an effect and judge whether a preprocessing choice has changed it. They are not a substitute for formal inferential analysis.

#### Safeguards, dependencies, and maintenance

NESTApp includes safeguards that prevent technically incompatible configurations while preserving investigator control over methodological decisions. Pipelines are checked before execution to identify missing dependencies, incompatible step ordering, or required parameters that have not been specified. For example, steps requiring continuous data cannot be placed after epoching. A saved pipeline that refers to an unavailable step still opens, so that it can be inspected or adapted, and is held back from execution with the missing dependency identified. Determining whether a given preprocessing approach is appropriate for a particular dataset remains the responsibility of the investigator.

NESTApp also includes mechanisms that help keep results stable as its underlying software dependencies evolve. Minimum plugin versions are specified where required, and an installation diagnostic reports the installed MATLAB, EEGLAB, and plugin versions. Where outputs depend on a specific implementation, such as the AARATEP template, the corresponding software release is fixed to an identified version rather than automatically updated. Automated testing is additionally used during NESTApp development to identify changes that could alter processing behaviour.

#### Demonstration

NESTApp processed 29 TMS-EEG recordings from 16 participants, acquired at 5 kHz over 63 channels, using the 32-step pipeline assembled in Fig. 2 and reported in Fig. 4. This analysis was intended to illustrate the implementation of the pipeline, its quality-control procedures, and the outputs generated by NESTApp, rather than to evaluate neurophysiological outcomes. The recordings were acquired in a study approved by the Research Ethics Board of The Royal’s Institute of Mental Health Research (REB 2019071) (Watson et al., 2023), all participants gave written informed consent, and the work was carried out in accordance with the Declaration of Helsinki.

Twenty-seven recordings completed. Two were stopped at TMS pulse detection and are reported as such rather than contributing partial output: a failed file does not stop the batch, and their input paths are written to a re-run list. Across the 27, 61 of 63 channels were retained with 6.2 *±* 3.2 interpolated, 71.6 *±* 16.3 of 76.1 *±* 17.0 epochs were retained, and 7.0 *±* 2.6 of 53.8 *±* 3.2 independent components were removed across the two rounds (mean *±* SD). Figure 7 shows one recording before and after.

**Figure 7.**
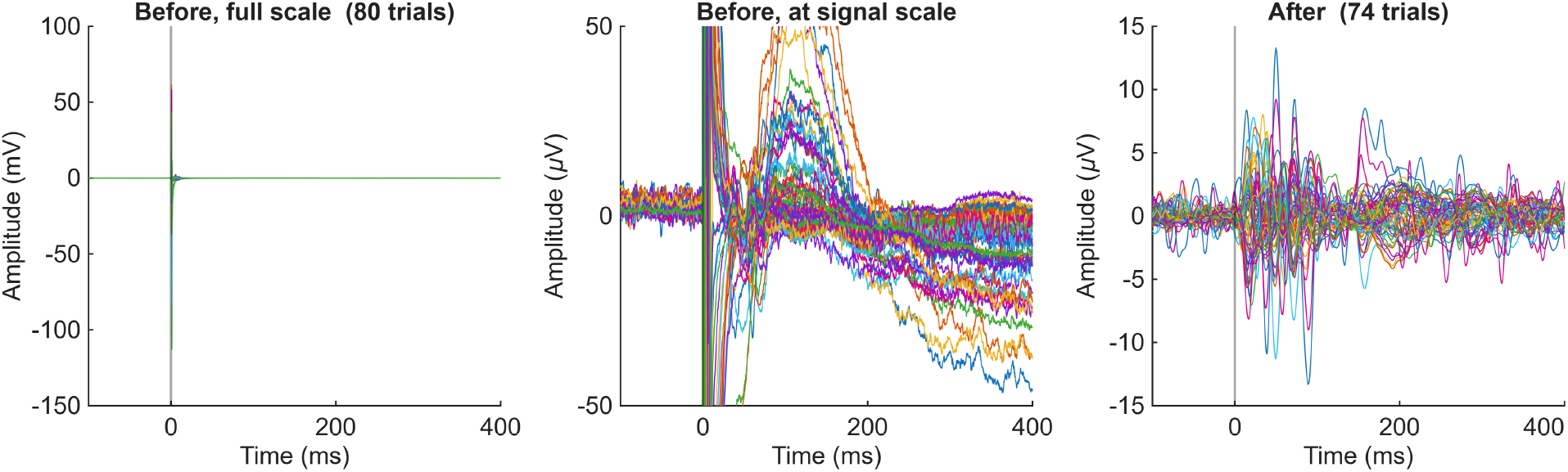
One recording before and after preprocessing. All 61 scalp channels of a single session, averaged over trials. **Left:** the recording with no artifact handling, at the scale the stimulation artifact demands. The pulse reaches 115 mV, four orders of magnitude above the response being sought, and nothing else in the recording is visible beside it. **Centre:** the same trace on a scale where an evoked response would show. What appears instead is the decay artifact and the slow drift that follow the pulse, still tens of microvolts at 300 ms. **Right:** the same recording through the 32-step pipeline, on which the evoked response is resolved within *±* 15 *μ*V. Note that each panel carries its own vertical scale. Both arms were executed by the same engine on the same input, so the comparison is between pipelines and not between tools; the left and centre panels are the minimum under which a TMS-evoked potential can be plotted at all, being load, channel selection, pulse detection, epoching, and baseline correction, with no artifact handling of any kind. Six trials of 80 were rejected by the full pipeline.

Three quality gates were configured, at load, after epoching, and before saving, with thresholds chosen to be plausible rather than tuned. They returned a spread rather than a uniform pass: 24 recordings passed every gate, one was marginal, and two failed. One recording is instructive: it was already marginal after epoching, at 42 trials, and fell below the minimum only by the end of the run, finishing at 39. A gate at only one of those points would have missed it or caught it too late to show when it went wrong. Figure 3 shows the verdicts for the batch.

## Limitations

The application requires MATLAB and the associated toolboxes listed in Table 1, which may limit accessibility for researchers without the necessary licences. In addition, although the automatically generated methods description documents the NESTApp version, processing steps, and associated parameters, it does not currently include the versions of EEGLAB and individual plugins used during preprocessing. This information is available through the installation diagnostic and is automatically captured when processing errors occur, but incorporating it directly into the processing record would further strengthen reproducibility.

Further validation of the application is also warranted. Although NESTApp implements established functions from EEGLAB and associated toolboxes, we have not systematically evaluated whether pipelines implemented through NESTApp produce identical outputs to equivalent pipelines executed using independently written scripts. Moreover, the current demonstration was conducted by the development team using data from a single cohort and acquisition environment. Evaluation across different TMS-EEG acquisition protocols, datasets, and independent users will therefore be important to establish the generalisability and usability of the application.

Finally, NESTApp is designed to support preprocessing rather than downstream neurophysiological analyses such as time-frequency, connectivity, or source modelling. Processed data are exported in standard EEGLAB format, so these analyses can be carried out with existing specialised tools. The group-level outputs currently provided within the *Explore* interface are intended primarily for visualisation and quality assessment and should not be interpreted as inferential statistical analyses.

## Conclusion

NESTApp provides an accessible framework for applying the same TMS-EEG preprocessing pipeline across users and datasets. Rather than introducing a new preprocessing algorithm or prescribing a specific pipeline, it integrates established tools within a flexible graphical environment while systematically documenting how preprocessing decisions are applied to each recording. By linking pipeline implementation, quality control, and reporting, NESTApp facilitates consistent processing across cohorts and improves the traceability of the resulting TMS-EEG measures. Its ability to save and share preprocessing workflows further allows them to be reapplied unchanged, without requiring users to write or modify analysis code. Together, these features may help reduce technical barriers to TMS-EEG preprocessing while promoting greater transparency and reproducibility across studies. Future development will extend this framework toward adaptive preprocessing approaches, including data-driven optimisation of parameters at the individual level.

## Data Availability

The 29 TMS-EEG recordings used in the Demonstration were acquired for an unrelated study and are not a scientific result of this work; they appear here to show what the application produces. They are available from the corresponding authors on reasonable request.

https://github.com/arefpz/nestapp

https://doi.org/10.5281/zenodo.23020967

## Code availability

NESTApp v2.2.0 is available at https://github.com/arefpz/nestappx under the GNU General Public License v3.0 or later, with documentation on the same repository. The release described here is archived at https://doi.org/10.5281/zenodo.23020967.

## Author Contributions

Conceptualization: A.P., S.T., J.L. Software: W.D. (lead), A.P. Validation: W.D. Visualization: W.D. Writing – original draft: A.P. Writing – review & editing: all authors. Supervision: S.T., J.L. Funding acquisition: S.T.

## Use of generative AI

Anthropic’s Claude was used in developing NESTApp, to draft and refactor source code and tests under the authors’ direction. It was not used to design the application or to choose the preprocessing methods it offers. All code was reviewed and tested by the authors, who take full responsibility for the software.

## References

Ablin, P., Cardoso, J.F., Gramfort, A., 2018. Faster independent component analysis by preconditioning with Hessian approximations. IEEE Transactions on Signal Processing 66, 4040–4049. doi:10.1109/tsp.2018.2844203.

Atluri, S., Frehlich, M., Mei, Y., Garcia Dominguez, L., Rogasch, N.C., Wong, W., Daskalakis, Z.J., Farzan, F., 2016. TMSEEG: a MATLAB-based graphical user interface for processing electrophysiological signals during transcranial magnetic stimulation. Frontiers in Neural Circuits 10, 78. doi:10.3389/fncir.2016.00078.

Barker, A.T., Jalinous, R., Freeston, I.L., 1985. Non-invasive magnetic stimulation of human motor cortex. The Lancet 325, 1106–1107. doi:10.1016/s0140-6736(85)92413-4.

Beck, M.M., Heyl, M., Mejer, L., Vinding, M.C., Christiansen, L., Tomasevic, L., Siebner, H.R., 2024. Methodological choices matter: A systematic comparison of TMS–EEG studies targeting the primary motor cortex. Human Brain Mapping 45, e70048. doi:10.1002/hbm.70048.

Bigdely-Shamlo, N., Mullen, T., Kothe, C., Su, K.M., Robbins, K.A., 2015. The PREP pipeline: standardized preprocessing for large-scale EEG analysis. Frontiers in Neuroinformatics 9, 16. doi:10.3389/fninf.2015.00016.

Casarotto, S., Romero Lauro, L.J., Bellina, V., Casali, A.G., Rosanova, M., Pigorini, A., Defendi, S., Mariotti, M., Massimini, M., 2010. EEG responses to TMS are sensitive to changes in the perturbation parameters and repeatable over time. PLoS ONE 5, e10281. doi:10.1371/journal.pone.0010281.

Cline, C.C., Lucas, M.V., Sun, Y., Menezes, M., Etkin, A., 2021. Advanced artifact removal for automated TMS-EEG data processing, in: 2021 10th International IEEE/EMBS Conference on Neural Engineering (NER), IEEE. pp. 1039–1042. doi:10.1109/ner49283.2021.9441147.

Corthout, E., Hallett, M., Cowey, A., 2011. TMS-induced blinking assessed with high-speed video: optical disruption of visual perception. Experimental Brain Research 210, 243–250. doi:10.1007/s00221-011-2626-y.

Cousineau, D., 2005. Confidence intervals in within-subject designs: A simpler solution to Loftus and Masson’s method. Tutorials in Quantitative Methods for Psychology 1, 42–45. doi:10.20982/tqmp.01.1.p042.

Delorme, A., 2023. EEG is better left alone. Scientific Reports 13, 2372. doi:10.1038/s41598-023-27528-0.

Delorme, A., Makeig, S., 2004. EEGLAB: an open source toolbox for analysis of single-trial EEG dynamics including independent component analysis. Journal of Neuroscience Methods 134, 9–21. doi:10.1016/j.jneumeth.2003.10.009.

Esposito, R., Bortoletto, M., Miniussi, C., 2020. Integrating TMS, EEG, and MRI as an approach for studying brain connectivity. The Neuroscientist 26, 471–486. doi:10.1177/1073858420916452.

Gabard-Durnam, L.J., Mendez Leal, A.S., Wilkinson, C.L., Levin, A.R., 2018. The Harvard automated processing pipeline for electroencephalography (HAPPE): standardized processing software for developmental and high-artifact data. Frontiers in Neuroscience 12, 97. doi:10.3389/fnins.2018.00097.

Gorjan, D., Gramann, K., De Pauw, K., Marusic, U., 2022. Removal of movement-induced EEG artifacts: current state of the art and guidelines. Journal of Neural Engineering 19, 011004. doi:10.1088/1741-2552/ac542c.

Gramfort, A., Luessi, M., Larson, E., Engemann, D.A., Strohmeier, D., Brodbeck, C., Goj, R., Jas, M., Brooks, T., Parkkonen, L., et al., 2013. MEG and EEG data analysis with MNE-Python. Frontiers in Neuroscience 7, 267. doi:10.3389/fnins.2013.00267.

Hernandez-Pavon, J.C., Veniero, D., Bergmann, T.O., Belardinelli, P., Bortoletto, M., Casarotto, S., Casula, E.P., Farzan, F., Fecchio, M., Julkunen, P., et al., 2023. TMS combined with EEG: Recommendations and open issues for data collection and analysis. Brain Stimulation 16, 567–593. doi:10.1016/j.brs.2023.02.009.

Hui, J., Tremblay, S., Daskalakis, Z.J., 2019. The current and future potential of transcranial magnetic stimulation with electroencephalography in psychiatry. Clinical Pharmacology & Therapeutics 106, 734–746. doi:10.1002/cpt.1541.

Hyvarinen, A., 1999. Fast and robust fixed-point algorithms for independent component analysis. IEEE Transactions on Neural Networks 10, 626–634. doi:10.1109/72.761722.

Ilmoniemi, R.J., Hernandez-Pavon, J.C., Mäkelä, N.N., Metsomaa, J., Mutanen, T.P., Stenroos, M., Sarvas, J., 2015. Dealing with artifacts in TMS-evoked EEG, in: 2015 37th Annual International Conference of the IEEE Engineering in Medicine and Biology Society (EMBC), IEEE. pp. 230–233. doi:10.1109/embc.2015.7318342.

Klug, M., Jeung, S., Wunderlich, A., Gehrke, L., Protzak, J., Djebbara, Z., Argubi-Wollesen, A., Wollesen, B., Gramann, K., 2022. The BeMoBIL pipeline for automated analyses of multimodal mobile brain and body imaging data. bioRxiv doi:10.1101/2022.09.29.510051. preprint.

Kothe, C.A., Makeig, S., 2013. BCILAB: a platform for brain-computer interface development. Journal of Neural Engineering 10, 056014. doi:10.1088/1741-2560/10/5/056014.

Lioumis, P., Kičić, D., Savolainen, P., Mäkelä, J.P., Kähkönen, S., 2009. Reproducibility of TMS—evoked EEG responses. Human Brain Mapping 30, 1387–1396. doi:10.1002/hbm.20608.

Morey, R.D., 2008. Confidence intervals from normalized data: A correction to Cousineau (2005). Tutorials in Quantitative Methods for Psychology 4, 61–64. doi:10.20982/tqmp.04.2.p061.

Mullen, T., 2012. CleanLine EEGLAB plugin. Neuroimaging Informatics Tools and Resources Clearinghouse (NITRC). RRID:SCR_002233.

Mutanen, T., Mäki, H., Ilmoniemi, R.J., 2013. The effect of stimulus parameters on TMS–EEG muscle artifacts. Brain Stimulation 6, 371–376. doi:10.1016/j.brs.2012.07.005.

Mutanen, T.P., Biabani, M., Sarvas, J., Ilmoniemi, R.J., Rogasch, N.C., 2020. Source-based artifact-rejection techniques available in TESA, an open-source TMS–EEG toolbox. Brain Stimulation 13, 1349–1351. doi:10.1016/j.brs.2020.06.079.

Oostenveld, R., Fries, P., Maris, E., Schoffelen, J.M., 2011. FieldTrip: open source software for advanced analysis of MEG, EEG, and invasive electrophysiological data. Computational Intelligence and Neuroscience 2011, 156869. doi:10.1155/2011/156869.

Pion-Tonachini, L., Kreutz-Delgado, K., Makeig, S., 2019. ICLabel: An automated electroencephalographic independent component classifier, dataset, and website. NeuroImage 198, 181–197. doi:10.1016/j.neuroimage.2019.05.026.

Rogasch, N.C., Fitzgerald, P.B., 2013. Assessing cortical network properties using TMS–EEG. Human Brain Mapping 34, 1652–1669. doi:10.1002/hbm.22016.

Rogasch, N.C., Sullivan, C., Thomson, R.H., Rose, N.S., Bailey, N.W., Fitzgerald, P.B., Farzan, F., Hernandez-Pavon, J.C., 2017. Analysing concurrent transcranial magnetic stimulation and electroencephalographic data: A review and introduction to the open-source TESA software. NeuroImage 147, 934–951. doi:10.1016/j.neuroimage.2016.10.031.

Rogasch, N.C., Thomson, R.H., Daskalakis, Z.J., Fitzgerald, P.B., 2013. Short-latency artifacts associated with concurrent TMS–EEG. Brain Stimulation 6, 868–876. doi:10.1016/j.brs.2013.04.004.

Saari, J., Kallioniemi, E., Tarvainen, M., Julkunen, P., 2018. Oscillatory TMS-EEG-responses as a measure of the cortical excitability threshold. IEEE Transactions on Neural Systems and Rehabilitation Engineering 26, 383–391. doi:10.1109/tnsre.2017.2779135.

Sazgar, M., Young, M.G., 2019. EEG artifacts, in: Absolute Epilepsy and EEG Rotation Review: Essentials for Trainees. Springer, pp. 149–162. doi:10.1007/978-3-030-03511-2\_8.

Sohn, Y., Voller, B., Dimyan, M., Gibson, A.S.C., Hanakawa, T., Leon-Sarmiento, F., Jung, H., Hallett, M., 2004. Cortical control of voluntary blinking: a transcranial magnetic stimulation study. Clinical Neurophysiology 115, 341–347. doi:10.1016/j.clinph.2003.10.035.

Tadel, F., Baillet, S., Mosher, J.C., Pantazis, D., Leahy, R.M., 2011. Brainstorm: A user-friendly application for MEG/EEG analysis. Computational Intelligence and Neuroscience 2011, 879716. doi:10.1155/2011/879716.

Tremblay, S., Rogasch, N.C., Premoli, I., Blumberger, D.M., Casarotto, S., Chen, R., Di Lazzaro, V., Farzan, F., Ferrarelli, F., Fitzgerald, P.B., et al., 2019. Clinical utility and prospective of TMS–EEG. Clinical Neurophysiology 130, 802–844. doi:10.1016/j.clinph.2019.01.001.

Veniero, D., Bortoletto, M., Miniussi, C., 2009. TMS-EEG co-registration: on TMS-induced artifact. Clinical Neurophysiology 120, 1392–1399. doi:10.1016/j.clinph.2009.04.023.

Watson, M., Chaves, A.R., Gebara, A., Desforges, M., Broomfield, A., Landry, N., et al., 2023. A naturalistic study comparing the efficacy of unilateral and bilateral sequential theta burst stimulation in treating major depression – the U-B-D study protocol. BMC Psychiatry 23, 739. doi:10.1186/s12888-023-05243-4.

Widmann, A., Schröger, E., Maess, B., 2015. Digital filter design for electrophysiological data—a practical approach. Journal of Neuroscience Methods 250, 34–46. doi:10.1016/j.jneumeth.2014.08.002.

Wu, W., Keller, C.J., Rogasch, N.C., Longwell, P., Shpigel, E., Rolle, C.E., Etkin, A., 2018. ARTIST: A fully automated artifact rejection algorithm for single-pulse TMS-EEG data. Human Brain Mapping 39, 1607–1625. doi:10.1002/hbm.23938.

