## Supplementary Information for "NESTApp: A GUI-Based MATLAB Application for Auditable TMS-EEG Preprocessing with EEGLAB"

Table S1 lists every step the application offers. The built-in templates are described in the main text.

Table S1: **Every preprocessing step offered by NESTApp.**

| Step | Provided by | What it does |
|---|---|---|
| **Import & Session** | | |
| Load Data | EEGLAB | Reads the selected recording, in EEGLAB, BrainVision, Neuroscan or Curry format |
| Load Channel Location | EEGLAB | Attaches electrode coordinates from a location file, and can validate the montage |
| Choose Data Set | EEGLAB | Retrieves a dataset held earlier in the run, to branch back without reloading |
| Save New Set | EEGLAB | Writes the current dataset, named from the input file and a suffix |
| **Channels** | | |
| Remove Unneeded Channels | EEGLAB | Keeps or drops named channels; by default the ear-reference electrodes |
| Detect Bad Channels (RANSAC) | clean_rawdata | Reconstructs each channel from a random subset of the others and drops those that correlate poorly with their own reconstruction |
| Detect Bad Channels (TESA) | TESA | Flags channels by robust deviation or a Wiener estimate, then interpolates, drops or blanks them |
| Remove Bad Channels | EEGLAB | Rejects channels whose kurtosis, probability or spectrum lies too many standard deviations from the mean |
| Remove Bad Channels (manual) | EEGLAB | Draws the scalp topography so channels can be ringed by hand and dropped |
| Interactive Channel Reject (TESA) | TESA | Opens a window for inspecting channels and marking them bad |
| Interpolate Channels | EEGLAB | Restores removed channels by spherical spline interpolation from their neighbours |
| **TMS Artifact** | | |
| Find TMS Pulses (TESA) | TESA | Detects pulses as large deflections in one reference electrode; handles paired-pulse and repetitive protocols |
| Find TMS Pulses (AARATEP) | AARATEP | Detects pulses from the artifact across channels, requiring a quarter of them to cross threshold together |
| Remove TMS Artifacts (TESA) | TESA | Blanks the pulse window in each epoch and records the cut for later interpolation |
| Fix TMS Pulse (TESA) | TESA | Realigns epochs to the steepest deflection, where the recorded marker mistimes the pulse |
| Interpolate Missing Data (TESA) | TESA | Fills the blanked window by linear or cubic interpolation |
| Interpolate Missing Data (AR-Blend) | AARATEP | Fills the blanked window by autoregressive extrapolation from each side, crossfaded across the gap |
| Remove Decay Artifact | AARATEP | Fits an exponential decay to the early post-pulse samples and subtracts it, per trial if asked |
| Fit Artifact Model (TESA) | TESA | Models the early decay from the recording's own traces and subtracts the fit |
| Find Artifacts EDM (TESA) | TESA | Finds muscle-artifact components by enhanced deflation, in the window just after the pulse |
| SSP-SIR | TESA | Projects the data away from the principal components of the early muscle artifact |
| Source-Informed Sensor Cleaning (SOUND) | TESA | Suppresses sensor noise using source-space constraints |
| **Reference & Resample** | | |
| Re-Reference | EEGLAB | Re-references to an electrode, a list of electrodes, or the common average |
| Re-Sample | EEGLAB | Changes the sampling rate, anti-alias filtering before it downsamples |
| **Detrend** | | |
| Detrend Epoch | EEGLAB | Removes a polynomial trend from each channel and epoch |
| Detrend (TESA) | TESA | Fits a linear, exponential or double-exponential trend over a window and subtracts it |
| Robust Detrend (TESA) | TESA | Fits a polynomial with outlying samples excluded, so the response itself cannot drag the fit |
| Robust Demean (TESA) | TESA | Subtracts a mean computed with outlying samples excluded: a level shift rather than a slope |

*Table S1 continued*

| Step | Provided by | What it does |
|---|---|---|
| **Filter** | | |
| Frequency Filter | firfilt | Hamming-windowed sinc FIR filter: high-pass, low-pass, band-pass or notch |
| Frequency Filter (TESA) | TESA | Zero-phase Butterworth filter: band-pass, band-stop, high-pass or low-pass |
| Frequency Filter (CleanLine) | CleanLine | Removes line noise and its harmonics by sinusoidal regression rather than by a notch |
| Modified Bandpass Filter (TESA) | TESA | Filters with autoregressive extrapolation across the blanked window, so the filter cannot ring off the artifact edge |
| Median Filter 1D | TESA | Replaces each sample with a neighbourhood median, removing spikes without smoothing the waveform |
| **Automatic Cleaning** | | |
| Clean Artifacts | clean_rawdata | Runs artifact subspace reconstruction over continuous data: flat and noisy channels, drifts, bursts, and windows it could not repair |
| Automatic Cleaning Data | clean_rawdata | Removes flat channels, noisy channels and burst artifacts from continuous data |
| Automatic Continuous Rejection | EEGLAB | Rejects stretches of continuous data whose spectral power exceeds a threshold |
| **Epochs & Trials** | | |
| Epoching | EEGLAB | Cuts fixed-length epochs time-locked to the chosen event |
| Remove Baseline | EEGLAB | Subtracts the mean of a baseline window from every trial |
| Remove Bad Epochs | EEGLAB | Rejects epochs by peak-to-peak amplitude and by joint probability |
| Remove Bad Trials | EEGLAB | Flags epochs by joint probability, then opens the rejection window for review before removing them |
| **ICA** | | |
| Run ICA (FastICA) | FastICA | Decomposes the data into independent components by FastICA |
| Run ICA (Infomax) | EEGLAB | Decomposes by infomax, extended by default: the family ICLabel was trained on |
| Run ICA (Picard) | PICARD | Decomposes by Picard, which reaches the infomax solution faster |
| Run ICA (TESA) | TESA | Runs FastICA through TESA, for use with its own component selector |
| Label ICA Components | ICLabel | Gives each component a probability of being brain, muscle, eye, heart, line noise or channel noise |
| Flag ICA Components for Rejection | ICLabel | Flags components whose label probability falls in a given range |
| Remove Flagged ICA Components | EEGLAB | Subtracts the flagged components from the data |
| Remove ICA Components (TESA) | TESA | Detects and removes TMS-evoked muscle, blink, eye-movement and muscle components by its own criteria |
| **TEP Analysis** | | |
| Extract TEP (TESA) | TESA | Averages across trials, over a region of interest or as global mean field amplitude |
| Find TEP Peaks (TESA) | TESA | Searches given windows for the expected positive and negative peaks |
| TEP Peak Output | TESA | Returns amplitude or area, and latency, for the peaks found |
| **Utilities & Inspection** | | |
| Quality Gate | nestapp | Measures the data against numeric thresholds and records Pass, Marginal or Fail; changes nothing |
| Visualize EEG Data | EEGLAB | Opens a scrolling plot and pauses the run for inspection |
| Manual Command | nestapp | Runs MATLAB code at this point, with the EEGLAB variables in scope |

A step is offered in the interface only when the toolbox providing it is installed. *nestapp* marks a step implemented in the application itself. The descriptions say what a step does to the data; its parameters, their units and their valid ranges are declared in the application and shown beside the step. Four steps are interactive and so cannot run under parallel processing: the two manual channel rejectors, *Remove Bad Trials*, and *Visualize EEG Data*.
